# Air pollution exposure and mitochondrial DNA copy number in the UK Biobank

**DOI:** 10.1101/2022.04.18.22273996

**Authors:** Yun Soo Hong, Stephanie L. Battle, Daniela Puiu, Wen Shi, Nathan Pankratz, Di Zhao, Dan E. Arking, Eliseo Guallar

## Abstract

**Background:** Low levels of mitochondrial DNA copy number (mtDNA-CN) are a biomarker of mitochondrial dysfunction often induced by oxidative stress. Air pollution is a pervasive source of oxidative stress, but the association between air pollution exposure and mtDNA-CN is inconclusive.

**Objective:** We evaluated the association between long-term exposure to PM ≤ 10 µm diameter (PM_10_) and nitrogen dioxide (NO_2_) with mtDNA-CN in 195,196 adults in the UK Biobank study.

**Methods:** Annual average PM_10_ and NO_2_ concentrations were estimated using separate land-use regression models for years 2007 and 2010. mtDNA-CN was measured in blood samples collected between 2007 and 2010 and was calculated as the ratio of mitochondrial coverage to the nuclear genome coverage derived from whole-genome sequencing in 195,196 UK Biobank participants. We used standardized values (Z-scores) after log-transformation as the mtDNA-CN metric.

**Results:** The median (interquartile range) annual average concentrations of PM_10_ and NO_2_ were 21.8 (20.3–23.6) and 28.9 (23.7–35.1) µg/m^3^ in 2007, and 16.2 (15.5–17.1) and 27.8 (22.7–32.4) µg/m^3^ in 2010. An increase of 10 µg/m^3^ in annual average PM_10_ and NO_2_ exposure was associated with an adjusted difference in mtDNA-CN of −0.089 (95% confidence interval; −0.090, −0.087) and −0.018 (−0.018, −0.017), respectively. The associations persisted for lags of up to 3 years. PM_2.5-10_ was also inversely associated with mtDNA-CN.

**Conclusions:** In this large-scale study, long-term exposure to PM_10_ and NO_2_ were inversely associated with mtDNA-CN. These findings suggest that oxidative stress-induced mitochondrial dysfunction, reflected by reduced mtDNA-CN, may be an additional mechanism mediating the health effects of air pollution.

## INTRODUCTION

Ambient air pollution is associated with a wide range of acute and chronic health outcomes, including all-cause mortality, cardiovascular and pulmonary morbidity and mortality, cognitive decline, and increased hospital and emergency department visits.^1–8^ Exposure to ambient particulate matter (PM) is the 5^th^ leading risk factor for mortality worldwide, causing an estimated 4.2 million deaths and a loss of over 103 million disability-adjusted life years in 2015.^2^ The adverse health effects of air pollution are mediated through oxidative stress, alterations in innate and adaptive immunity, inflammation, and epigenetic changes.^9–12^

Mitochondria are essential to energy production, generating over 90% of the cell’s energy in the form of adenosine triphosphate (ATP).^13^ Mitochondria also play a major role in the regulation of metabolism and multiple homeostatic, inflammatory, and apoptotic signaling pathways.^14^ Each cell has tens to thousands of mitochondria, and each mitochondrion has 2–10 copies of the mitochondrial chromosome, a circular, double-stranded, haploid DNA strand that encodes 37 genes involved in oxidative phosphorylation or in assembling amino acids into functional proteins.^15^ Mitochondrial DNA (mtDNA) is susceptible to oxidative stress caused by excessive reactive oxygen species (ROS) because of its proximity to the mitochondrial membrane where oxidation products are generated and because of its inefficient protection and repair mechanisms.^16^ Moreover, damaged mtDNA induces excess ROS production, which may further aggravate and accumulate mtDNA damage.^17, 18^ Decreased mtDNA copy number (mtDNA-CN) is a marker of mitochondrial dysfunction that can be measured in peripheral blood and is associated with increased all-cause mortality,^19, 20^ cardiovascular disease,^21–24^ and chronic kidney disease (CKD).^25^

While air pollution is a pervasive source of oxidative stress, the association between exposure to air pollution and mtDNA-CN has been inconclusive.^26–35^ Among 2,758 healthy women in the Nurses’ Health Study, higher 12-month average particulate matter ≤ 2.5 µm in diameter (PM_2.5_) was associated with lower mtDNA-CN.^26^ On the other hand, occupational exposures to PM and respirable dust were associated with higher mtDNA-CN.^31, 32^ Previous studies are difficult to compare because of small sample sizes,^27, 29, 33^ inadequate control of potential confounders,^28, 29^ wide variations in exposure concentration and composition,^32, 34^ possible differences between short- and long-term effects of air pollution, and heterogeneity in exposure assessment and mtDNA-CN measurement.

We therefore evaluated the association between long-term exposure to air pollution (PM_10_, PM_2.5_, PM_2.5-10_, NO_2_, and NO) with mtDNA-CN in over 190,000 adult men and women from the UK Biobank study.

## METHODS

### Study population

The UK Biobank is a prospective cohort study of approximately 500,000 adults 40 to 69 years of age recruited across the United Kingdom (UK) from 2006 to 2010 via mail based on the UK National Health Services registration.^36^ Among 199,945 UK Biobank participants with whole genome sequencing (WGS) data available, we excluded individuals who had their blood samples collected in year 2006 (n = 170) or who had abnormal blood cell counts (n = 377; Supplementary Figure 1). We further excluded participants with missing values on body mass index (BMI; n = 824) or information on genetic relatedness (n = 165). The final study population was 198,409 (89,138 men and 109,271 women). Analyses for each specific pollutant were further restricted to participants with information on the concentration of each air pollutant (Supplementary Table 1).

All participants provided written informed consent prior to participation. The scientific protocol and operational procedures of the UK Biobank study were approved by the North West – Haydock Research Ethics Committee in the UK. The Institutional Review Board of the Johns Hopkins School of Medicine gave ethical approval for this work.

### Measurement of air pollutant concentrations

We used data on air pollution estimates provided by the Small Area Health Statistics Unit as part of the BioSHaRE-EU Environmental Determinants of Health project (http://www.sahsu.org/). The data were available for 2007 and 2010. For each participant, we used their residential address at the time of the UK Biobank visit to estimate annual average concentrations of each air pollutant.

The estimates of PM_10_ and NO_2_ for 2007 were derived from European Union (EU)-wide air pollution maps based on a land-use regression (LUR) model for Europe, which incorporated satellite-derived air pollution estimates, with a resolution of 100 × 100 m.^37^ The adjusted R^2^ across the UK for PM_10_ and NO_2_ were 0.57 and 0.64, respectively.

The estimates of air pollutants for 2010 were modeled using a LUR model developed as part of the European Study of Cohorts for Air Pollution Effects (ESCAPE).^38, 39^ The R^2^ of training models for PM_2.5_, PM_2.5-10_, and NO in the UK were 0.35–0.60, 0.68–0.79, and 0.83–0.89, respectively. The accuracy of particulate matter estimates was unclear for areas beyond the range of 400 km from Greater London and, therefore, air pollution concentrations were coded as missing for all addresses beyond this range.

### Measurement of mitochondrial DNA copy number

To estimate mtDNA-CN in UK Biobank participants, we ran our newly developed MitoHPC pipeline^40^ on WGS data accessed through the UK Biobank Research Analysis Platform which contains WGS information from peripheral blood in 199,945 UK Biobank participants. We first obtained the number of mapped reads through Samtools (idxstats command) which we used to calculate the nuclear genome and mitochondrial genome coverage. Nuclear genome coverage was calculated as the product of the total number of aligned reads and the read length divided by the estimated human genome size (3.03 G bp). Mitochondrial genome coverage was calculated as the product of the number of reads aligned to mitochondria and the read length divided by mitochondrial DNA size (16,569 bp). The read length was 151 bp. A raw mtDNA-CN was then calculated as 2 × mitochondrial coverage / nuclear genome coverage. mtDNA-CN was then log-transformed and standardized by subtracting the mean and dividing by the standard deviation (SD). The mtDNA-CN metric in this study thus represents SD units (Z-scores).

For mtDNA-CN measurement, WGS performs substantially better than qPCR or whole exome sequencing (WES).^41^ However, as WGS was available only in a subset of the UK Biobank population, we performed a sensitivity analysis using a separate mtDNA-CN metric generated using a combination of WES data and genotyping array probe intensity data available in the full UK Biobank cohort.^42^ In brief, with WES data from 49,997 individuals, we generated residuals from a linear regression model of mitochondrial chromosome read counts adjusted for total DNA and potential technical artifacts. We then used rank-transformed mitochondrial SNP probe intensities and generated 250 principal components (PCs) from autosomal nuclear probes by randomly sampling 5% of probes (n ∼ 19,500 probes) from odd chromosomes that were on both UKBelieve and Axiom arrays. For each array type, all mitochondrial SNP probes (UKBelieve, n = 181; Axiom, n = 244) and 250 PCs were regressed on the residuals derived from the linear regression model using the WES results. Coefficients from each model were subsequently used to generate the predicted values in all UK Biobank participants. The estimates were also standardized by subtracting the mean and dividing by the SD (Z-scores). After applying the same exclusion criteria as the main analysis, 378,607 participants (174,055 men and 204,552 women) were included in the sensitivity analysis.

### Measurement of other covariates

Study participants visited one of 22 assessment centers where they completed a touch-screen questionnaire and a computer-assisted interview and provided blood, urine, and saliva samples. Physical measures, including height, weight, and blood pressure, were also taken. All assessments were administered according to a standardized procedure. Participants provided information on age, sex, race / ethnicity, average total household income before tax (<£18,000, £18,000–£30,999, £31,000–£51,999, £52,000–£100,000, and >£100,000), and education (categorized as less than college, college or university degree, professional degree, or none of the above). Smoking status was categorized as never, former, and current smokers. Alcohol intake was categorized as never, former, and current drinkers. Physical activity was measured by self-report using the adapted International Physical Activity Questionnaire (IPAQ) Short Form,^43, 44^ which was converted into total physical activity in metabolic equivalent task (MET) minutes per week by intensity (walking, moderate, and vigorous physical activity).^45^ Total METs were then categorized as low, moderate, and high physical activity based on the IPAQ guideline.^43^

Body mass index (BMI) was calculated as weight in kg divided by height in m squared. Blood pressure was measured using an automated blood pressure monitor (Omron 705 IT, OMRON Healthcare Europe BV). Hypertension was defined as a self-reported physician’s diagnosis of hypertension, a self-reported use of antihypertensive medication, or a measured systolic blood pressure ≥ 140 mmHg, or diastolic blood pressure ≥90 mmHg. Prevalence of myocardial infarction (MI)^46^ and stroke were defined by an algorithm developed by the UK Biobank.^47^ Cardiovascular disease was defined as the presence of either MI or stroke.

Complete blood count, differential white blood cell (WBC) counts (neutrophils, lymphocytes, monocytes, eosinophils, and basophils), and reticulocytes were measured on fresh samples using an a Beckman automated hematology analyzer.^48^ Serum glucose levels were measured by hexokinase analysis, total cholesterol levels by CHO-POD method, low-density lipoprotein (LDL) cholesterol levels by enzymatic protective selection analysis, and triglyceride levels by GPO-POD method (Beckman Coulter AU5800, UK). Hemoglobin A1c (HbA1c) was measured in frozen packed red blood cells using high-performance liquid chromatography (Bio-Rad Variant II Turbo analyzer, Bio-Rad Laboratories, US). Details of the laboratory measurements can be found in the UK Biobank online showcase (http://ukbiobank.ac.uk). Diabetes was defined as a self-reported physician’s diagnosis of diabetes, a self-reported use of antidiabetic medication, or a measured HbA1c ≥ 6.5%. Hyperlipidemia was defined as a self-reported use of lipid-lowering medication, or a measured total cholesterol ≥200 mg/dl or triglycerides ≥150 mg/dl.

### Statistical analysis

For the main analysis, we used the average concentrations of PM_10_ and NO_2_ in 2007 and 2010 as the exposure and mtDNA-CN derived from blood samples collected in 2007–2010 as the outcome. That is, air pollution measured in 2007 was used as exposure for mtDNA-CN derived from samples collected in 2007–2010 (lags of 0, 1, 2, and 3 years), and air pollution measured in 2010 was used as exposure for mtDNA-CN derived from samples collected in 2010 (lag 0) to ensure that the exposure was measured either in the same year or before the outcome (Supplementary Table 1). All measurements were combined into a single dataset for the main analyses, but we also conducted separate analyses by year of air-pollution exposure and blood sample collection. For PM_2.5_, PM_2.5-10_, and NO concentrations with mtDNA-CN measures, we used only air pollution and mtDNA-CN measures from 2010.

We used linear regression models with progressive degrees of adjustment to estimate the association between a 10 µg/m^3^ increase in air pollutant exposure and mtDNA-CN using covariates measured at the time of the UK Biobank visit. Model 1 was adjusted for age, sex, self-reported ethnic background, year of air pollution measurement, and year of blood collection. Model 2 was further adjusted for average annual income, education level, smoking, alcohol intake, physical activity, BMI, and history of hypertension, diabetes, hyperlipidemia, and cardiovascular disease. Finally, Model 3 was further adjusted for blood cell counts. Since participants could be genetically related, and also since participants who had blood samples collected in 2010 were included twice in the analysis (using air pollution exposure levels in 2007 and in 2010), we used sandwich covariance matrix estimation (sandwich package in R) using a degrees of freedom-based correction to correct the standard errors for these sources of dependence among observations.

In addition to modeling air pollution as a linear term, we compared the differences in mtDNA-CN by quintiles of air pollutant concentrations using the lowest quintile as the reference, and we modeled air pollutant concentrations as restricted cubic splines with knots at the 5^th^, 27.5^th^, 50^th^, 72.5^th^, and 95^th^ percentiles to provide a smooth and flexible description of the dose-response relationship between air pollution and mtDNA-CN.

We performed stratified analyses to identify subgroups that may potentially be more susceptible using pre-specified subgroups: age (<60 or ≥ 60), sex, self-reported ethnic background (White, Black, Asian, other), BMI (underweight, normal, overweight, or obese), smoking status (never, former, or current), alcohol intake (never, former, or current), physical activity (low, moderate, or high), hypertension, and diabetes. Finally, we performed sensitivity analysis using the mtDNA-CN metric derived from the WES and mitochondrial SNP probe intensities in the full UK Biobank cohort. For this analysis, because the metric was validated in unrelated White individual, we excluded non-White individuals and who had genetically related individuals (used.in.pca.calculation variable provided by the UK Biobank) in the study. All statistical analyses were performed using R software version 4.1.2.

## RESULTS

The mean age (SD) of the 198,409 study participants (89,138 men and 109,271 women) was 56.5 (8.1) years (Table 1). Compared to participants in the lower quintiles of mtDNA-CN, those in the higher quintiles of mtDNA-CN were more likely to be younger and female, to have higher average annual income, to be never or former smokers and physically active, and to have lower body mass index, systolic blood pressure, serum glucose, LDL cholesterol, triglyceride levels, and lower prevalence of hypertension, diabetes, and cardiovascular disease. The median (interquartile range) annual average concentrations of PM_10_ and NO_2_ were 21.8 (20.3–23.6) and 28.9 (23.7–35.0) µg/m^3^, respectively, in 2007, and 16.2 (15.5–17.1) and 27.8 (22.7–32.4) µg/m^3^, respectively, in 2010 (Supplementary Table 1). The characteristics of participants included in the PM_10_ and the NO_2_ analyses were similar.

**Table 1.** Participant characteristics by quintiles of mitochondrial DNA copy number.

|  | **Overall**<br>**(n=198,409)** | **Quintile 1**<br>**(n=43,955)** | **Quintile 2**<br>**(n=39,585)** | **Quintile 3**<br>**(n=38,521)** | **Quintile 4**<br>**(n=38,788)** | **Quintile 5**<br>**(n=37,560)** |
| --- | --- | --- | --- | --- | --- | --- |
| **mtDNA-CN\*** | -0.05<br>(-4.84, 7.02) | -1.20<br>(-4.84, -0.78) | -0.48<br>(-0.78, -0.26) | 0.01<br>(-0.26, 0.21) | 0.51<br>(0.21, 0.80) | 1.32<br>(0.80, 7.02) |
| **Age (years)** | 56.5 (8.1) | 57.5 (8.1) | 56.8 (8.1) | 56.5 (8.1) | 56.0 (8.1) | 55.4 (8.0) |
| **Men (%)** | 44.9 | 49.3 | 46.2 | 44.4 | 42.8 | 41.2 |
| **Self-reported ethnic background (%)** |  |  |  |  |  |  |
| White | 94.1 | 94.5 | 94.4 | 94.5 | 94.2 | 92.8 |
| Black | 1.5 | 0.9 | 1.0 | 1.3 | 1.6 | 2.8 |
| Asian | 2.4 | 2.6 | 2.5 | 2.2 | 2.3 | 2.2 |
| Other | 1.6 | 1.5 | 1.7 | 1.5 | 1.5 | 1.7 |
| **Average income (%)** |  |  |  |  |  |  |
| < £18,000 | 18.9 | 21.8 | 19.4 | 18.8 | 17.7 | 16.3 |
| £18,000 – £30,999 | 21.6 | 22.4 | 21.9 | 21.4 | 21.4 | 20.6 |
| £31,000 – £51,999 | 22.6 | 21.1 | 22.3 | 22.9 | 23.2 | 23.7 |
| £52,000 – £100,000 | 17.8 | 15.4 | 17.2 | 17.8 | 18.7 | 20.2 |
| > £100,000 | 4.7 | 4.0 | 4.6 | 4.8 | 5.0 | 5.5 |
| **Education (%)** |  |  |  |  |  |  |
| Less than college | 37.8 | 36.8 | 37.8 | 38.2 | 38.2 | 38.4 |
| College or university degree | 33.1 | 30.6 | 32.5 | 32.9 | 34.1 | 35.8 |
| Professional degree | 11.6 | 12.1 | 11.5 | 11.7 | 11.4 | 11.2 |
| **Smoking (%)** |  |  |  |  |  |  |
| Never | 55.1 | 52.7 | 54.6 | 55.0 | 56.0 | 57.2 |
| Former | 34.8 | 35.4 | 34.4 | 35.0 | 34.4 | 34.6 |
| Current | 9.7 | 11.3 | 10.4 | 9.5 | 9.1 | 7.8 |
| **Alcohol intake (%)** |  |  |  |  |  |  |
| Never | 4.3 | 4.6 | 4.3 | 4.3 | 4.1 | 4.1 |
| Former | 3.4 | 4.0 | 3.6 | 3.3 | 3.2 | 2.9 |
| Current | 92.0 | 91.0 | 91.8 | 92.2 | 92.5 | 92.8 |
| **Physical activity (%)** |  |  |  |  |  |  |
| Low | 15.0 | 15.9 | 15.2 | 14.8 | 14.9 | 14.2 |
| Moderate | 33.0 | 32.8 | 33.4 | 33.2 | 32.9 | 32.9 |
| High | 32.9 | 31.2 | 32.3 | 33.2 | 33.5 | 34.4 |
| **BMI (kg/m<sup>2</sup>)** | 27.4 (4.7) | 28.0 (5.1) | 27.7 (4.8) | 27.4 (4.6) | 27.1 (4.5) | 26.6 (4.4) |
| **SBP (mmHg)** | 137.6 (18.4) | 139.3 (18.7) | 138.1 (18.5) | 137.5 (18.4) | 136.9 (18.3) | 136.0 (18.1) |
| **Glucose (mg/dL)** | 92.2 (22.0) | 93.8 (24.8) | 92.7 (22.9) | 92.0 (21.0) | 91.5 (20.9) | 90.7 (19.2) |
| **Total cholesterol (mg/dL)** | 220.3 (43.9) | 219.9 (45.3) | 220.9 (44.1) | 220.8 (43.7) | 220.5 (43.2) | 219.2 (43.0) |
| **LDL cholesterol (mg/dL)** | 137.5 (33.4) | 137.7 (34.4) | 138.2 (33.5) | 138.0 (33.3) | 137.5 (32.9) | 136.2 (32.7) |
| **Triglyceride (mg/dL)<sup>§</sup>** | 130.9 (96.5) | 139.6 (103.0) | 136.0 (99.2) | 132.0 (96.4) | 127.2 (93.7) | 118.5 (86.7) |
| **Hypertension (%)** | 52.9 | 58.3 | 54.6 | 52.7 | 50.5 | 47.6 |
| **Diabetes (%)** | 5.8 | 7.6 | 6.0 | 5.6 | 5.0 | 4.3 |
| **Hyperlipidemia (%)** | 80.6 | 82.5 | 81.7 | 81.1 | 79.9 | 77.3 |
| **Prevalent MI (%)** | 1.6 | 2.1 | 1.7 | 1.5 | 1.4 | 1.2 |
| **Prevalent stroke (%)** | 1.4 | 1.8 | 1.4 | 1.4 | 1.2 | 1.2 |
| **Prevalent CVD (%)** | 2.9 | 3.8 | 2.9 | 2.8 | 2.5 | 2.3 |

Abbreviations: BMI, body mass index; CVD, cardiovascular disease; LDL, low-density lipoprotein; MI, myocardial infarction; mtDNA-CN, mitochondrial DNA copy number; NO<sub>2</sub>, nitrogen dioxide; PM, particulate matter; SBP, systolic blood pressure.
\* Median (range); <sup>§</sup>Median (interquartile range).

In the fully-adjusted model, an increase in PM_10_ of 10 µg/m^3^ was associated with a difference in mtDNA-CN of −0.088 (95% confidence interval [CI] −0.090, −0.087; Table 2). For NO_2_, the corresponding difference in mtDNA-CN was −0.018 (−0.018, −0.017). Compared to participants in the lowest quintile of air pollutant exposure, the difference in mtDNA-CN among those in the highest quintile of PM_10_ exposure was −0.090 (−0.091, −0.089; *P* for trend < 0.001) and among those in the highest quintile of NO_2_ exposure it was −0.033 (−0.046, −0.019; *P* for trend < 0.001). The inverse trend for PM_10_ with mtDNA-CN was also evident in spline regression analyses (Figure 1). For NO_2_, there was an inverse association between NO_2_ and mtDNA-CN at concentrations above 30 µg/m^3^. The associations did not materially change after adjusting for seasonality and additional hematologic parameters (not shown). In sensitivity analyses, the associations were similar when we used the mtDNA-CN metric generated using WES and mitochondrial SNP probe intensities (Supplementary Table 2).

**Figure 1.**
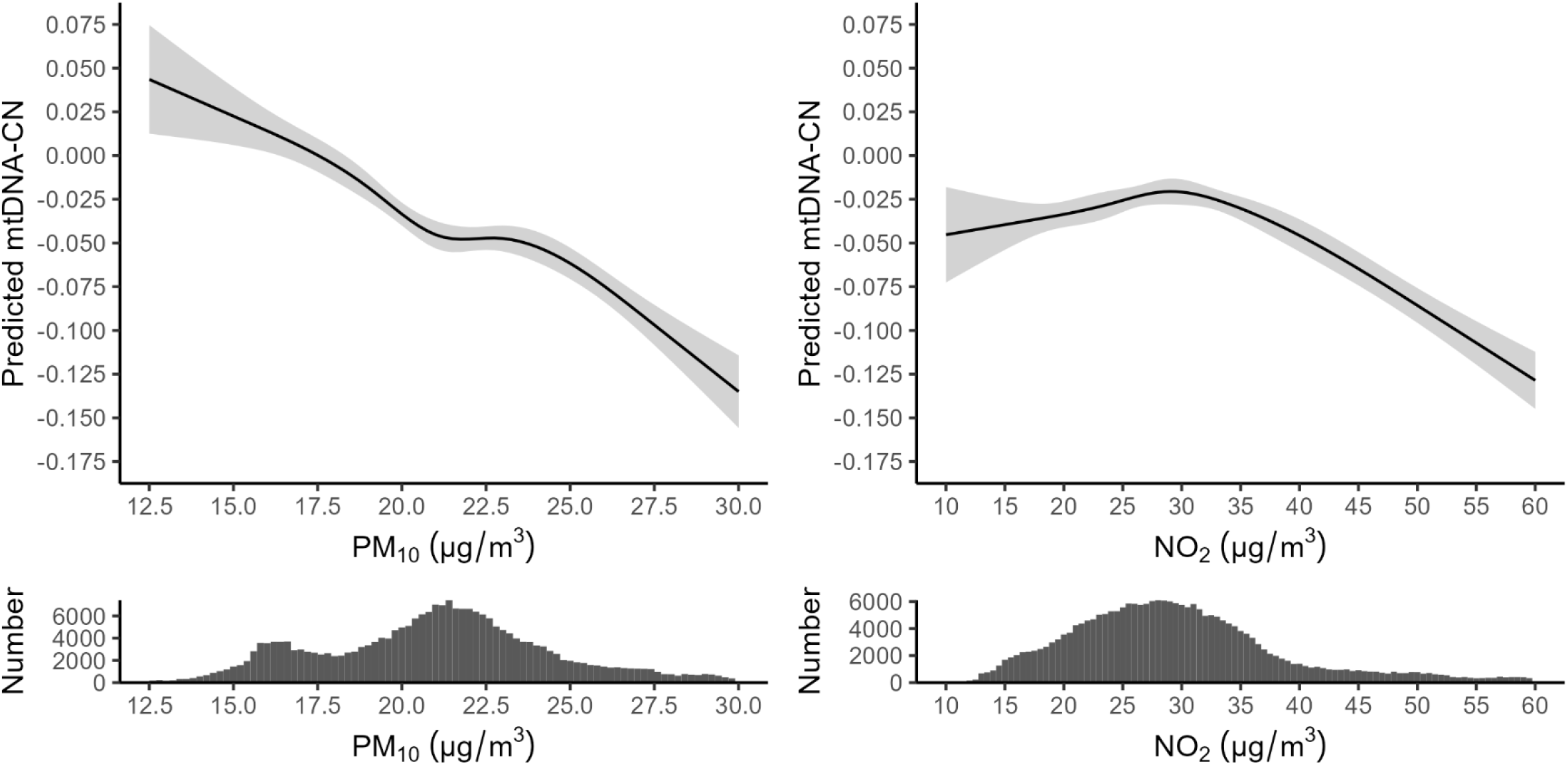
Average mitochondrial DNA copy number levels by PM_10_ and NO_2_ concentrations. The curves represent estimated mitochondrial DNA copy number levels (solid line) and their 95% confidence intervals (gray area) by PM_10_ and NO_2_ concentrations based on fully adjusted regression models using restricted cubic splines to model air pollutants with knots at the 5^th^, 27.5^th^, 50^th^, 72.5^th^, and 95^th^ percentiles of its distribution. The spline regression model was adjusted for age, sex, self-reported ethnic background, year of air pollution measurement, year of blood collection, average annual income, education level, smoking, alcohol intake, physical activity, body mass index, history of hypertension, diabetes, hyperlipidemia, and cardiovascular disease, and blood cell counts (red blood cell, neutrophils, lymphocytes, monocytes, eosinophils, basophils, and platelets).

**Table 2.** Average difference in mitochondrial DNA copy number (95% confidence interval) associated with a 10 µg/m^3^ increase in PM_10_ and NO_2_ and by quintile of air pollutant.

| | Linear<br>(per 10 $\mu\text{g}/\text{m}^3$<br>increase) | Quintile 1 | Quintile 2 | Quintile 3 | Quintile 4 | Quintile 5 | P trend |
| --- | --- | --- | --- | --- | --- | --- | --- |
| **$\text{PM}_{10}$** |  | 11.8, 17.8<br>(n = 48,125) | 17.8, 20.4<br>(n = 48,219) | 20.4, 21.9<br>(n = 47,851) | 21.9, 23.7<br>(n = 48,084) | 23.7, 36.6<br>(n = 47,904) |  |
| **Model 1** | -0.107<br>(-0.108, -0.105) | Reference | -0.038<br>(-0.048, -0.029) | -0.071<br>(-0.089, -0.053) | -0.082<br>(-0.098, -0.066) | -0.109<br>(-0.123, -0.096) | < 0.001 |
| **Model 2** | -0.102<br>(-0.104, -0.101) | Reference | -0.035<br>(-0.046, -0.025) | -0.058<br>(-0.073, -0.044) | -0.067<br>(-0.080, -0.054) | -0.100<br>(-0.111, -0.089) | < 0.001 |
| **Model 3** | -0.088<br>(-0.090, -0.087) | Reference | -0.038<br>(-0.042, -0.033) | -0.054<br>(-0.058, -0.050) | -0.059<br>(-0.062, -0.055) | -0.090<br>(-0.091, -0.089) | < 0.001 |
| **$\text{NO}_2$** |  | 7.3, 22.2<br>(n = 48,199) | 22.2, 26.7<br>(n = 48,078) | 26.7, 30.8<br>(n = 48,193) | 30.8, 36.0<br>(n = 48,062) | 36.0, 128.0<br>(n = 48,082) |  |
| **Model 1** | -0.023<br>(-0.023, -0.023) | Reference | -0.009<br>(-0.017, -0.000) | -0.021<br>(-0.031, -0.012) | -0.023<br>(-0.026, -0.020) | -0.058<br>(-0.073, -0.043) | < 0.001 |
| **Model 2** | -0.021<br>(-0.021, -0.020) | Reference | 0.007<br>(-0.003, 0.016) | 0.000<br>(-0.007, 0.007) | -0.002<br>(-0.006, 0.003) | -0.043<br>(-0.060, -0.026) | < 0.001 |
| **Model 3** | -0.018<br>(-0.018, -0.017) | Reference | 0.012<br>(0.010, 0.014) | 0.014<br>(0.012, 0.016) | 0.010<br>(0.008, 0.013) | -0.033<br>(-0.046, -0.019) | < 0.001 |

Abbreviations: $\text{NO}_2$ , nitrogen dioxide; PM, particulate matter.
Model 1: Adjusted for age, sex, self-reported ethnic background, year of air pollution measurement, and year of blood collection; Model 2: Model 1 + average annual income, education level, smoking, alcohol intake, physical activity, body mass index, and history of hypertension, diabetes, hyperlipidemia, and cardiovascular disease; Model 3: Model 2 + cell counts (red blood cells, neutrophil, lymphocytes, basophils, eosinophils, monocytes, and platelets).

In the analysis using air pollutant concentrations and mtDNA-CN measured in the concurrent year (Lag 0), an increase of 10 µg/m^3^ in PM_10_ and NO_2_ was associated with a difference in mtDNA-CN of −0.358 (−0.415, −0.300) and −0.060 (−0.075, −0.045), respectively, in 2007, and with a difference of −0.053 (−0.100, −0.006) and −0.007 (−0.019, 0.005), respectively, in 2010 (Table 3). For PM_10_ and NO_2_ concentrations from 2007 and mtDNA-CN measured in 2008–2010, analyses conducted separately by year of measurement of mtDNA-CN consistently showed an inverse association but did not show a clear trend with increasing lag (Table 3). For PM_2.5_, PM_2.5-10_, and NO concentrations and mtDNA-CN measured only in 2010, the fully-adjusted differences in mtDNA-CN associated with an increase of 10 µg/m^3^ were −0.040 (−0.135, 0.055), −0.127 (−0.226, −0.027), and −0.004 (−0.009, 0.002), respectively (Table 4).

**Table 3.** Average difference in mitochondrial DNA copy number associated with a 10 µg/m^3^ increase in PM_10_ and NO_2_ by lag of measurement between air pollution and mtDNA copy number (0, 1, 2, and 3 years).

| mtDNA copy number measured in | Air pollutant measured in 2007 |  |  |  | Air pollutant measured in 2010 |
| --- | --- | --- | --- | --- | --- |
|  | 2007 (Lag 0) | 2008 (Lag 1) | 2009 (Lag 2) | 2010 (Lag 3) | 2010 (Lag 0) |
| **$\text{PM}_{10}$** | Range: 13.0, 32.7<br>(n = 18,717) | Range: 12.2, 36.5<br>(n = 67,488) | Range: 11.8, 36.6<br>(n = 63,927) | Range: 12.8, 36.6<br>(n = 45,005) | Range: 11.8, 26.2<br>(n = 45,046) |
| **Model 1** | -0.365<br>(-0.424, -0.306) | -0.086<br>(-0.115, -0.056) | -0.102<br>(-0.129, -0.074) | -0.081<br>(-0.109, -0.054) | -0.076<br>(-0.124, -0.029) |
| **Model 2** | -0.331<br>(-0.394, -0.268) | -0.073<br>(-0.105, -0.041) | -0.077<br>(-0.107, -0.047) | -0.101<br>(-0.131, -0.071) | -0.078<br>(-0.129, -0.026) |
| **Model 3** | -0.358<br>(-0.415, -0.300) | -0.068<br>(-0.098, -0.039) | -0.061<br>(-0.088, -0.033) | -0.080<br>(-0.108, -0.052) | -0.053<br>(-0.100, -0.006) |
| **$\text{NO}_2$** | Range: 7.3, 102.0<br>(n = 18,720) | Range: 7.3, 111.3<br>(n = 67,609) | Range: 8.3, 128.3<br>(n = 64,153) | Range: 8.4, 117.9<br>(n = 45,066) | Range: 12.9, 97.7<br>(n = 45,066) |
| **Model 1** | -0.074<br>(-0.089, -0.059) | -0.017<br>(-0.026, -0.008) | -0.027<br>(-0.034, -0.020) | -0.014<br>(-0.021, -0.006) | -0.011<br>(-0.023, 0.001) |
| **Model 2** | -0.055<br>(-0.071, -0.039) | -0.005<br>(-0.015, 0.004) | -0.023<br>(-0.031, -0.015) | -0.019<br>(-0.027, -0.011) | -0.011<br>(-0.024, 0.002) |
| **Model 3** | -0.060<br>(-0.075, -0.045) | -0.006<br>(-0.015, 0.003) | -0.019<br>(-0.026, -0.012) | -0.014<br>(-0.022, -0.007) | -0.007<br>(-0.019, 0.005) |

Abbreviations: mtDNA, mitochondrial DNA; $\text{NO}_2$ , nitrogen dioxide; PM, particulate matter.
Model 1: Adjusted for age, sex, and self-reported ethnic background; Model 2: Model 1 + average annual income, education level, smoking, alcohol intake, physical activity, body mass index, and history of hypertension, diabetes, hyperlipidemia, and cardiovascular disease; Model 3: Model 2 + cell counts (red blood cells, neutrophil, lymphocytes, basophils, eosinophils, monocytes, and platelets).

**Table 4.** Average difference in mitochondrial DNA copy number associated with a 10 µg/m^3^ increase in PM_2.5_, PM_2.5-10_, and NO measured concurrently in 2010 (Lag 0).

|  |  |
| --- | --- |
| **$\text{PM}_{2.5}$** | Range: 8.2, 18.4<br>(n = 45,046) |
| **Model 1** | -0.073 (-0.168, 0.022) |
| **Model 2** | -0.052 (-0.155, 0.051) |
| **Model 3** | -0.040 (-0.135, 0.055) |
| **$\text{PM}_{2.5-10}$** | Range: 5.6, 12.2<br>(n = 45,046) |
| **Model 1** | -0.119 (-0.219, -0.019) |
| **Model 2** | -0.155 (-0.263, -0.046) |
| **Model 3** | -0.127 (-0.226, -0.027) |
| **NO** | Range: 19.7, 252.0<br>(n = 45,066) |
| **Model 1** | -0.005 (-0.010, 0.001) |
| **Model 2** | -0.003 (-0.010, 0.003) |
| **Model 3** | -0.004 (-0.009, 0.002) |

Abbreviations: mtDNA, mitochondrial DNA; NO, nitric oxide; PM, particulate matter.
Model 1: Adjusted for age, sex, and self-reported ethnic background; Model 2: Model 1 + average annual income, education level, smoking, alcohol intake, physical activity, body mass index, and history of hypertension, diabetes, hyperlipidemia, and cardiovascular disease; Model 3: Model 2 + cell counts (red blood cells, neutrophil, lymphocytes, basophils, eosinophils, monocytes, and platelets).

In stratified analysis, the associations between PM_10_ and mtDNA-CN were more pronounced in the higher BMI categories (*P* for interaction < 0.001), in current smokers (*P* for interaction < 0.001), and in participants with hypertension (*P* for interaction = 0.03) and with diabetes (*P* for interaction < 0.001; Figure 2). The associations were similar across other subgroups examined. The subgroup analysis for NO_2_ showed similar results.

**Figure 2.**
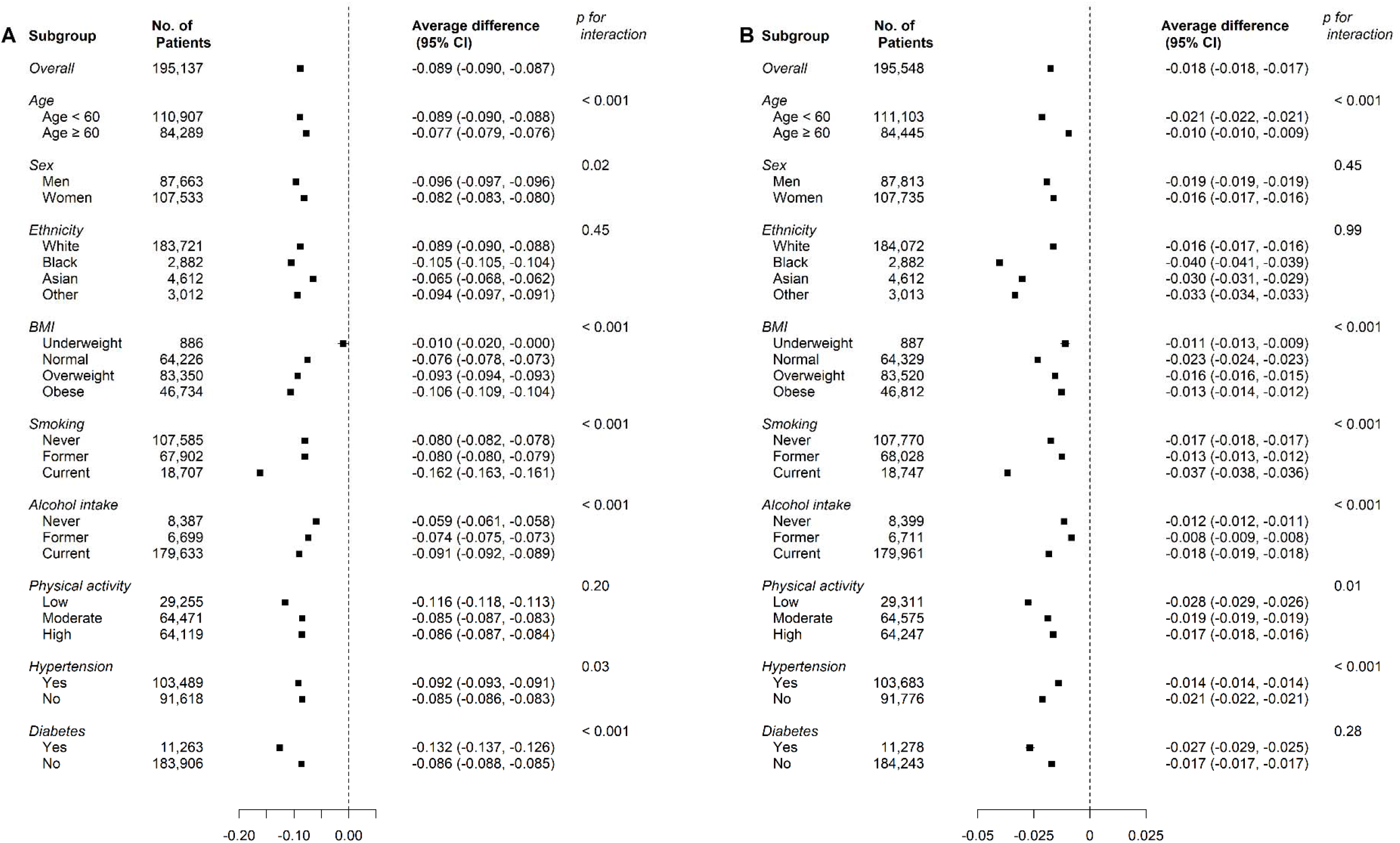
Average difference in mitochondrial DNA copy number associated with a 10 µg/m^3^ increase in PM_10_ (A) and NO_2_ (B) in subgroups. The figure displays the difference in mitochondrial DNA copy number associated with a 10 µg/m^3^ increase in PM_10_ (A) and NO_2_ (B) in models adjusted for age, sex, race, year of air pollution measurement, year of blood collection, average annual income, education level, smoking, alcohol intake, physical activity, body mass index, history of hypertension, diabetes, hyperlipidemia, and cardiovascular disease, and blood cell counts (red blood cell, neutrophils, lymphocytes, monocytes, eosinophils, basophils, and platelets). Pre-specified subgroups were age (<60 or ≥ 60), sex, self-reported ethnic background (White, Black, Asian, or Other), body mass index (underweight, normal, overweight, or obese), smoking status (never, former, or current), alcohol intake (never, former, or current), physical activity (low, moderate, or high), and presence of hypertension and diabetes, separately.

## DISCUSSION

In this large-scale population-based study, we found that higher concentrations of PM_10_ and NO_2_ were inversely associated with mtDNA-CN concentrations measured in blood. These findings support the hypothesis that oxidative stress-induced mitochondrial dysfunction, reflected by reduced mtDNA-CN, is a potential mechanism linking air pollution with various adverse health outcomes.

Air pollution is a major cause of mortality and global burden of disease.^1, 2^ Its health effects include increased mortality, hospital admissions and emergency department visits, and higher risk of pulmonary, cardiovascular, neurodegenerative, and other diseases.^3–8^ Exposure to air pollutants causes local inflammation and oxidative stress in the lung and generation of ROS, triggering the release of cytokines, a systemic inflammatory response, and higher levels of stress hormones due to sympathetic activation. Chronic exposure to air pollution can alter innate and adaptive immunity and cause epigenetic changes.^9, 10, 49–51^

mtDNA-CN fluctuates with changes in energy demands, oxidative stress, and various physiological and environmental conditions.^15, 52^ mtDNA is a particularly susceptible target of ROS-induced damage and mutation as it lacks protective histones and has limited repair capacity.^16^ mtDNA damage and mutation result in inefficient mitochondrial function, which initially triggers mtDNA replication and increases in mtDNA-CN to enhance energy supply and remove damage. Under prolonged exposure and chronic stress, however, mutations accumulate, mtDNA-CN decreases, and mitochondria become dysfunctional.^17^ Mitochondrial dysfunction is characterized by impaired oxidative phosphorylation and energy synthesis, increased ROS generation, disruption of calcium homeostasis and calcium signaling pathways, and alterations in metabolic, inflammatory, and apoptotic signaling pathways due to changes in the expression of associated nuclear genes.^10, 15^ mtDNA-CN measured in peripheral blood is a useful biomarker reflecting oxidative stress-mediated mitochondrial dysfunction, and it has been associated with adverse health outcomes,^15^ including all-cause mortality,^19, 20^ cardiovascular disease,^21–24^ and CKD.^25^

Air pollutants can have adverse effects on mitochondrial structure and function.^10^ First, PM can impair the mitochondrial respiratory chain, calcium regulation, and mitochondrial membrane potential, resulting in reduced ATP production.^53–56^ For example, exposure to diesel exhaust PM decreased mitochondrial respiratory activity and reduced ATP production in rats.^53^ In addition, PM increased mitochondrial permeability transition in other animal models^54, 55^ and decreased mitochondrial membrane potential in endothelial cells.^56^ Second, PM can promote mitochondrial ROS formation via downregulation or loss of function of antioxidant enzymes^57^ and activation of inflammatory and inhibition of anti-inflammatory pathways.^58^ Third, PM may induce changes in the expression of genes involved in the fission and fusion of mitochondria,^10^ leading to both an inhibition of mitochondrial fusion^56^ and an increase in the production of giant mitochondria by the fusion of multiple mitochondria.^59^ Lastly, PM may damage the integrity and the function of mtDNA through increased levels of ROS within mitochondria or DNA methylation.^60^ However, further studies are needed to understand the link between other air pollutants, such as NO_2_, and mitochondrial dysfunction.

Long-term exposure to air pollution was inversely associated with mtDNA-CN in other studies. In a study of 2,758 healthy women from the Nurses’ Health Study, an interquartile range (5.5 µg/m^3^) increase in annual average PM_2.5_ was associated with a difference of −0.07 (95% CI - 0.13, −0.01) units in mtDNA-CN.^26^ In addition, PM exposure in the past year was inversely associated with mtDNA-CN in two elderly populations^30, 34^ and in three small other studies evaluating the short-term effects of air pollution. ^27, 29, 32^ However, PM_2.5_ exposure in the past month and the past week were positively associated with mtDNA-CN in an elderly population,^30^ and 5-day to 28-day average exposures of black carbon were associated with higher mtDNA-CN.^35^

The results of previous studies are difficult to compare because many studies were small, restricted to occupational exposures or to a highly selected population, and with limited adjustment for potential confounders. Moreover, the durations and concentrations of exposure and the methods of mtDNA-CN assessment varied substantially across studies. Despite these limitations, and consistent with our findings, long-term (≥1 year) exposure to air pollutants was mostly associated with lower mtDNA-CN. Lower mtDNA-CN levels measured in peripheral blood may indicate a more systemic response to long-term air pollution exposure as blood-derived mtDNA-CN is positively associated with gene expression in other tissues including heart, uterus, and arteries.^61^

Evidence on the relationship between non-PM air pollutants and mtDNA-CN is relatively scarce. In our study, NO_2_ was inversely associated with mtDNA-CN at higher levels. Similarly, higher concentrations of nitrate (NO_3_) as a component of PM_2.5_ were associated with lower mtDNA-CN in a study of elderly men.^34^ Prenatal exposure of NO_2_ in the last month and third trimester of pregnancy was also inversely associated with placental mtDNA-CN.^62^ We, however, did not find any association between NO and mtDNA-CN. Oxidation of atmospheric nitrogen (N_2_) forms NO at high temperatures, which, due to its unstable property, reacts rapidly with oxygen to form NO_2_.^63^ Therefore, NO_2_ or nitrogen oxides (NO_x_), which includes both NO and NO_2_, may be a better metric to represent long-term outdoor exposures and associated health effects.

We have also evaluated the associations of PM_10_ and NO_2_ with mtDNA-CN measured with different time intervals (Lags 0–3 years). Regardless of the time interval, each air pollutant was inversely associated with mtDNA-CN but we did not see a clear trend by time interval. These results are difficult to interpret because, in our analysis, different lags include different participants, making it difficult to know if the differences are due to the lags or to the participants included in each lag. In addition, the association between air pollution and mtDNA-CN at lag 0 in 2007 year was much larger than in 2010. This difference may be due to declining levels of air pollution between 2007 and 2010, higher mtDNA-CN in 2010, and differences in participant characteristics. In addition, different modeling methods were used for air pollution estimates in 2007 and 2010. Our analysis needs to be replicated in different populations with a varied range of temperature exposures. In addition, longitudinal studies with multiple measurements of air pollution levels and mtDNA-CN in the same individuals are needed to better understand the response of mtDNA-CN to changes in environmental stressors.

Several limitations need to be considered in the interpretation of our findings. First, mtDNA-CN was measured on a single occasion which limited our ability to evaluate the change in mtDNA-CN over time by exposure concentration. Second, there are inherent limitations of using air pollution concentrations from prediction models. Only ambient concentrations of air pollutants were available and we were not able to consider personal exposure levels that may vary by activity patterns, workplace exposures, and indoor air pollution. The measurement error in the air pollution levels, however, is likely to be unrelated to mtDNA-CN and would have attenuated the association. Third, as an observational study, there may be unmeasured confounders that we could not account for. Finally, the number of non-White individuals in the UK Biobank was small. Therefore, further studies including diverse populations are needed to better understand variability by race and ethnicity group.

## CONCLUSIONS

In this large-scale population-based study, long-term exposure to PM_10_ and NO_2_ were inversely associated with mtDNA-CN. These findings suggest oxidative stress-induced mitochondrial dysfunction, reflected by reduced mtDNA-CN, as a potential mechanism between air pollution and various adverse health outcomes.

## Data Availability

All data used in the study is available as part of the UK Biobank.

https://www.ukbiobank.ac.uk/

## ACKNOWLEDGEMENTS

None.

## SUPPLEMENTARY MATERIALS

**Supplementary Table 1.**
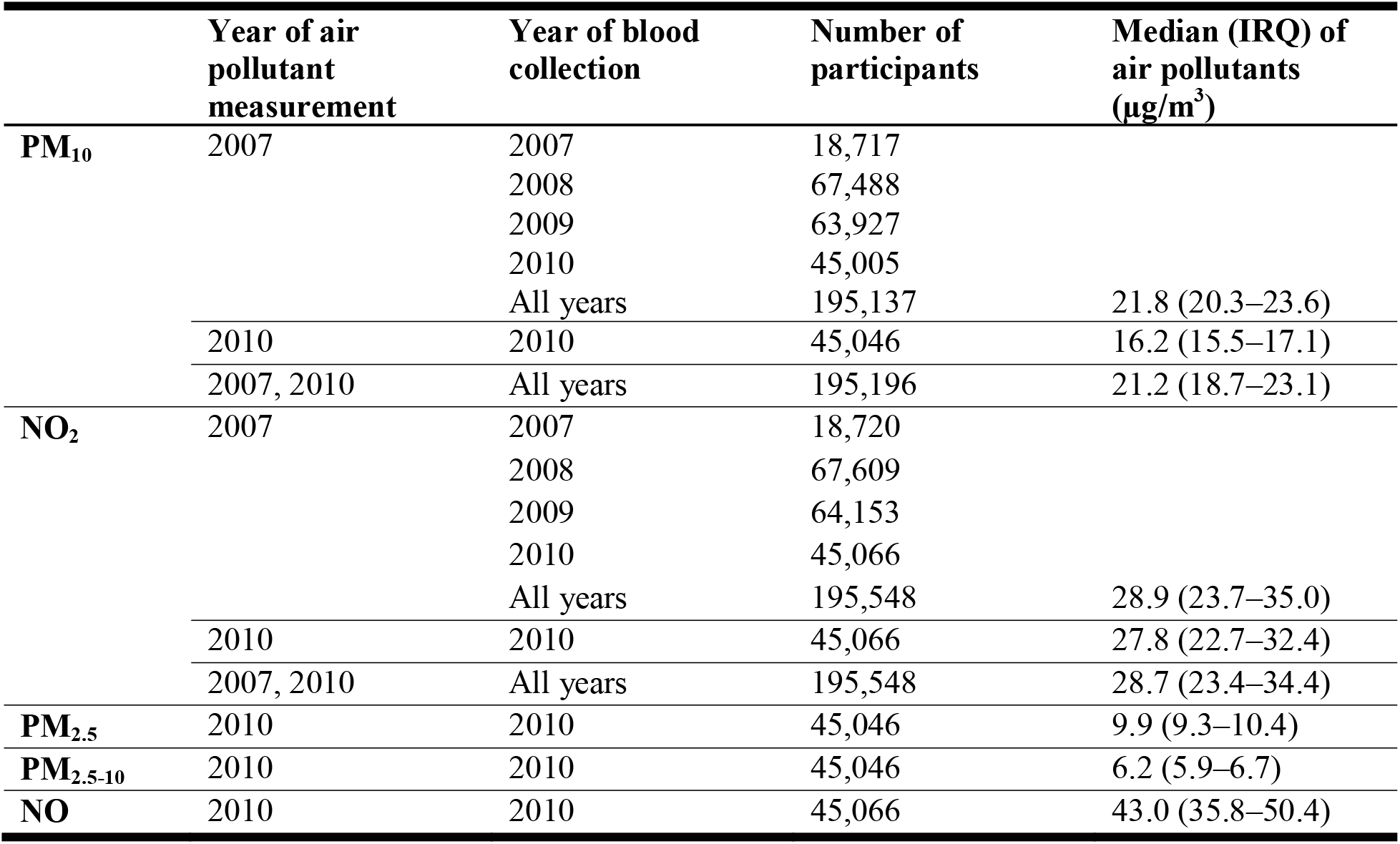
Description of data available on air pollutants and mitochondrial DNA copy number.

**Supplementary Table 2.** Average difference in mitochondrial DNA copy number (95% confidence interval) associated with a 10 µg/m^3^ increase in PM_10_ and NO_2_ and by quintile of air pollutant in the full UK Biobank.

|  | Linear | Quintile 1 | Quintile 2 | Quintile 3 | Quintile 4 | Quintile 5 | P trend |
| --- | --- | --- | --- | --- | --- | --- | --- |
| **$\text{PM}_{10}$** |  | 11.8, 18.3<br>(n = 87,306) | 18.3, 20.5<br>(n = 87,437) | 20.5, 21.9<br>(n = 87,259) | 21.9, 23.6<br>(n = 86,956) | 23.6, 36.6<br>(n = 86,766) |  |
| **Model 1** | -0.084<br>(-0.085, -0.083) | Reference | -0.026<br>(-0.038, -0.015) | -0.054<br>(-0.065, -0.042) | -0.054<br>(-0.066, -0.042) | -0.071<br>(-0.083, -0.059) | < 0.001 |
| **Model 2** | -0.079<br>(-0.080, -0.078) | Reference | -0.024<br>(-0.036, -0.011) | -0.048<br>(-0.060, -0.035) | -0.045<br>(-0.058, -0.032) | -0.064<br>(-0.077, -0.051) | < 0.001 |
| **Model 3** | -0.068<br>(-0.069, -0.067) | Reference | -0.024<br>(-0.036, -0.012) | -0.039<br>(-0.051, -0.027) | -0.035<br>(-0.047, -0.023) | -0.053<br>(-0.066, -0.041) | < 0.001 |
| **$\text{NO}_2$** |  | 7.0, 21.9<br>(n = 87,336) | 21.9, 26.4<br>(n = 87,488) | 26.4, 30.3<br>(n = 87,192) | 30.3, 35.4<br>(n = 87,385) | 35.4, 138.0<br>(n = 87,207) |  |
| **Model 1** | -0.024<br>(-0.024, -0.023) | Reference | -0.013<br>(-0.022, -0.004) | -0.028<br>(-0.037, -0.018) | -0.015<br>(-0.024, -0.005) | -0.058<br>(-0.068, -0.048) | < 0.001 |
| **Model 2** | -0.021<br>(-0.021, -0.020) | Reference | -0.006<br>(-0.016, 0.005) | -0.014<br>(-0.024, -0.004) | 0.004<br>(-0.006, 0.014) | -0.046<br>(-0.057, -0.035) | < 0.001 |
| **Model 3** | -0.019<br>(-0.019, -0.018) | Reference | 0.000<br>(-0.009, 0.010) | -0.003<br>(-0.013, 0.006) | 0.013<br>(0.004, 0.023) | -0.036<br>(-0.041, -0.026) | < 0.001 |

Abbreviations: $\text{NO}_2$ , nitrogen dioxide; PM, particulate matter.
In this analysis, mitochondrial DNA copy number was derived using whole exome sequencing (WES) and mitochondrial probe intensities to the full UK Biobank cohort.
Model 1: Adjusted for age, sex, self-reported ethnic background, year of air pollution measurement, and year of blood collection; Model 2: Model 1 + average annual income, education level, smoking, alcohol intake, physical activity, body mass index, and history of hypertension, diabetes, hyperlipidemia, and cardiovascular disease; Model 3: Model 2 + cell counts (red blood cells, neutrophil, lymphocytes, basophils, eosinophils, monocytes, and platelets).

**Supplementary Figure 1.**
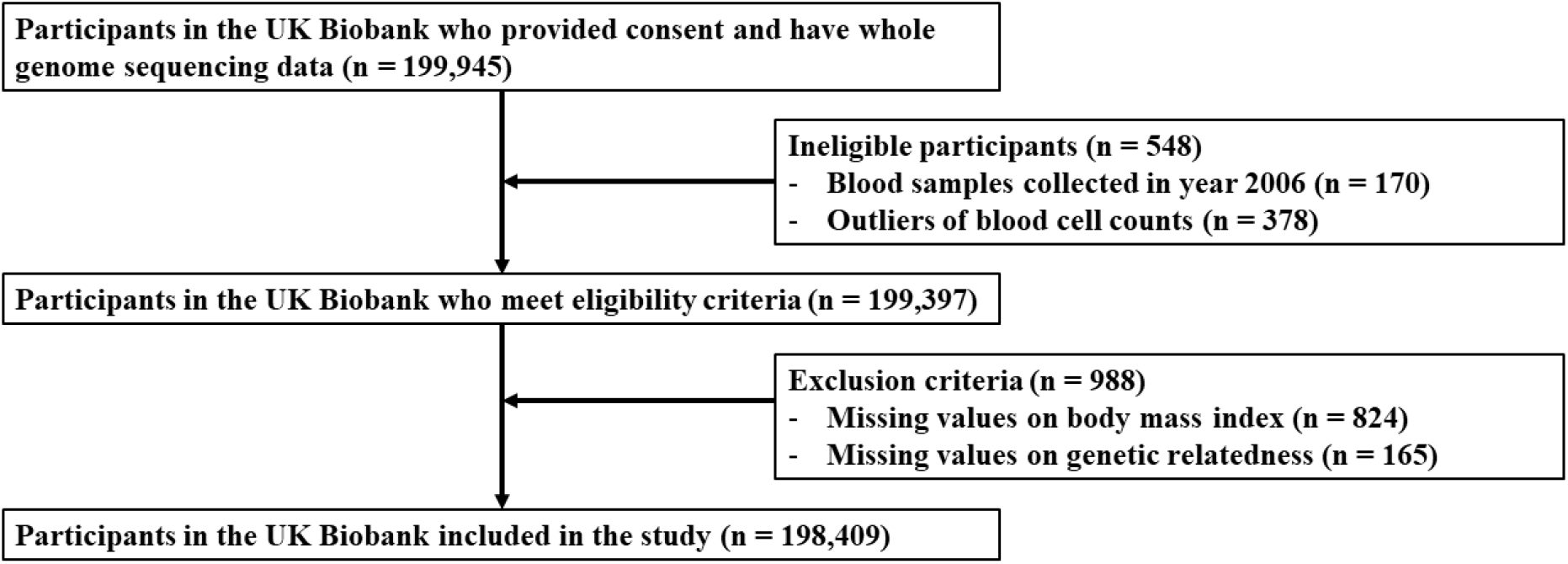
Flowchart of study participants.

